# Effects of Vascular Endothelial Growth Factor Family on Macular Pucker: *cis*-Mendelian Randomization and Colocalization Analyses

**DOI:** 10.1101/2024.12.01.24318223

**Authors:** Qi Gao, Jinglong Guo

## Abstract

**Background:** Vascular endothelial growth factor (VEGF) family and its receptors (VEGFR) could be implicated in macular pucker (MP) pathogenesis. We used *cis*-Mendelian randomization (*cis*-MR) and Bayesian colocalization with summary-level genome-wide association study (GWAS) data to explore causal relationships.

**Methods:** Genetic variants associated with soluble VEGFR2 (sVEGFR2), sVEGFR3, VEGF-A, VEGF-C, or VEGF-D levels were selected from a GWAS of protein quantitative trait loci with 35,559 Icelanders. MP GWAS (3,974 cases, 376,650 controls) was sourced from the FinnGen. We employed *cis*-MR using invariance-weighted median, supplemented by other methods. Bayesian colocalization validated *cis*-MR findings. Pleiotropy, reverse causality, and heterogeneity were assessed.

**Findings:** *Cis*-MR suggested that genetically predicted higher levels of sVEGFR2 were associated with reduced MP risk (Odds ratio (OR) 0.82, 95% confidence interval (CI) 0.75-0.89, P=9.20 × 10^-6^). Colocalization supported shared genetic variants in the *VEGFR2* gene region between sVEGFR2 and MP (posterior probability of hypothesis 4 (PPH_4_) =0.94), reinforcing sVEGFR2’s protective role in MP. Although *cis*-MR suggested an inverse relationship between sVEGFR3 levels and MP risk (OR 0.71, 95% CI 0.57-0.89, P=2.64 × 10^-3^), colocalization analysis did not confirm direct causality (PPH_4_=0.03). VEGF-A, VEGF-C and VEGF-D levels were not associated with MP risk in MR analyses. No evidence of pleiotropy, reverse causality, and heterogeneity was found across our MR analyses.

**Interpretation:** Our study suggested that sVEGFR2 has causally protective effects against MP and could serve as a potential drug target. Further research is necessary to elucidate its protective mechanisms and validate its translational implications.

## 1. Introduction

Macular pucker (MP), also known as epiretinal membrane, is a potentially sight-threatening vitreoretinal disease. It is characterized by a sheet of abnormal, thin, fibrotic tissue that grows on the surface of the macula that is responsible for detailed central vision. The prevalence of MP ranges from 7% to 11.8%, with aging being the primary nonmodifiable risk factor (1; 2; 3). Surgical intervention, such as vitrectomy with membrane peeling, is the current standard of care for symptomatic MP cases (1; 2; 3). However, pharmacological approaches targeting MP pathogenesis are still largely lacking, likely due to insufficient understanding of the causal factors involved in its pathogenesis (3; 4).

The pathogenesis of MP involves a complex interplay among various cells and acellular factors, encompassing angiogenesis, inflammation, and fibrosis at the vitreoretinal interface (3; 5; 6; 7). Numerous studies consistently found VEGF and VEGFR family factors in pathological tissues of MP patients, notably VEGF-A and its key receptor, VEGFR2, suggesting their potential role in MP pathogenesis (8; 9; 10; 11). VEGF-A predominantly binds to membrane-bound VEGFR2 (mVEGFR2) on endothelial cells of blood vessels, promoting crucial processes such as proliferation, migration, and survival, which are necessary for angiogenesis or vascular permeability (11; 12). Simultaneously, VEGF-C and VEGF-D regulate lymphangiogenesis through VEGFR3 on lymphatic endothelial cells (11; 12). VEGF-A and VEGF-B interact with VEGFR1, although their specific roles are less defined (11; 12). Notably, VEGF-A can cross-activate VEGFR3, whereas VEGF-C and VEGF-D can weakly bind to VEGFR2 (11; 12). Beyond their roles in angiogenesis and lymphangiogenesis, VEGF/VEGFR signaling is implicated in fibrosis and inflammation (3; 6; 11; 12). These pathways can be regulated by soluble forms of VEGFRs-such as sVEGFR1, sVEGFR2, and sVEGFR3-that act as decoy receptors for their respective VEGF molecules (13; 14; 15; 16; 17; 18). For example, studies have indicated that sVEGFR2 likely plays an important role in regulating angiogenesis in patients with macular edema, although the specific biological mechanisms were not characterized in these studies (19; 20). However, the complexity of these interactions complicates the identification of which member(s) of the VEGF/VEGFR family could be causally associated with pathogenesis of relevant diseases such as MP, especially in human studies.

Mendelian randomization (MR) is a method used to evaluate the potential causality of a modifiable exposure on an outcome by utilizing genetic variants as instrumental variables (21). Because genetic variants are randomly assorted during gamete formation and fixed at conception, MR estimates could minimize bias resulted from confounding factors or reverse causality (21). *Cis*-MR specifically refers to MR studies that use genetic variants from gene-restricted regions of pharmacological protein of interest, and it is typically employed for drug target validation (21; 22). Bayesian colocalization analysis evaluates whether two distinct traits are affected by the same causal variants, making it an important complementary analysis to appraise the validity and strength of a *cis*-MR estimate (23).

In this study, we investigated whether plasma levels of specific members of the VEGF/VEGFR family are causally associated with MP risk. We employed *cis*-MR and colocalization analyses and leveraged large-scale summary-level genome-wide association study (GWAS) of protein quantitative trait loci (pQTL) and MP to study this question.

## 2 Methods

### 2.1 Data source

Information of the data sources used in this study was summarized in Supplementary table 1. The study relied on summary-level GWAS data that have been available publicly, ethical approvals were obtained in all original studies. A GWAS of pQTL that included sVEGFR2, sVEGFR3, VEGF-A, VEGF-C, and VEGF-D was used to select genetic variants. The study included 35,559 participants of Icelanders and was available at https://www.decode.com/summarydata/ (24). In the original study, plasma proteins were measured using the SomaScan v4 platform with standard quality controls and adjusted for year of birth, sex, and year of sample collection (24). MP GWAS (N=3,974 cases, N=376,650 controls) was from the tenth release of FinnGen study, with cases being defined based on International Classification of Diseases (ICD-10) (https://www.finngen.fi/en/access_results). The original MP GWAS was adjusted for age, sex, and genetic principal components (25).

### 2.2 Selection of genetic instruments

A study of summary-level GWAS of pQTL involving 35,559 Icelandic participants was used to identify genetic variants associated with levels of sVEGFR2, sVEGFR3, VEGF-A, VEGF-C and VEGF-D, respectively (24).

For *cis*-MR, genetic variants near *sVEGFR2*, *sVEGFR3*, and *VEGF-C* genes within a 100 kilobase (kb) window were chosen, with a 200 kb window used for *VEGF-A* to ensure robust instrument strength. VEGF-D, of which gene located on the X chromosome, was not included in *cis*-MR. Variants were required to have a GWAS-correlated P-value < 5 x 10^-8^ and linkage disequilibrium (LD) r^2^ < 0.1 within a 100kb window based on the European 1000 Genome Project reference panel.

In univariable MR, stringent LD-clumping criteria were applied to select genome-wide variants associated with levels of sVEGFR2, sVEGFR3, VEGF-A, and VEGF-D, while VEGF-C variants did not meet these criteria and were excluded from univariable MR analyses. Variants were required to have a GWAS-correlated P-value < 5 x 10^-8^ and LD r^2^ < 0.001 within a 10,000kb window based on the European 1000 Genome Project reference panel. Genetic variants associated with MP were also selected under these criteria for reverse MR.

Variants with an F-statistic < 10 and minor allele frequency (MAF) < 0.01 were excluded. An F-statistic > 10 indicates robust instrument strength, calculated by F = R^2^ (N - K - 1)/K(1 - R^2^), where R^2^ is the proportion of variance in the exposure explained by genetic variants, K is the number of instruments, and N is the sample size (26). R^2^ estimates the proportion of variance in the phenotype explained by the genetic variant, R^2^ = 2 × (Beta/SD)^2^ × (MAF) × (1 − MAF), where Beta represents the per-allele effect size of the association between each variant and the phenotype, and SD denotes the standard deviation.

Functional variant annotation was performed using snpXplorer (27), which suggested no evidence of association with MP in our study. Detailed information on the genetic variants was provided in Supplementary table 1-10.

### 2.3 Univariable MR and *cis*-MR analyses

We performed MR analyses following the STROBE-MR guidelines (28). MR study should be conducted based on three core principles: (1) relevance, the genetic instruments should be significantly associated with the exposure; (2) independence, the genetic instruments should not be associated with any potential confounder and (3) exclusion restriction, the genetic instruments should not directly affect the outcome except via the way of exposure (21). Genetic correlations between exposure and outcome for each variant were harmonized by aligning effects alleles, with exclusion of palindromic variants. Prior to each MR analysis, MR Pleiotropy RESidual Sum and Outlier (MR-PRESSO) was applied to identify and remove potential outliers (29). The causal effects were assessed mainly using inverse variance-weighted (IVW) MR. This method assumes that outcomes are influenced only by the exposure, setting the intercept at zero (30). We also performed MR Egger and weighted median MR approaches. MR-Egger allows sensitivity tests to rule out potential bias induced by horizontal pleiotropy using the inclusion of the intercept in the regression analysis, and P < 0.05 was considered significant for the presence of horizontal pleiotropy (31). The weighted median MR provides a consistent estimate of the causal effect even when up to 50% of the information comes from invalid variants (i.e., those that violate the MR assumptions) (32). MR-PRESSO global test was also used to assess the presence of horizontal pleiotropy and P value < 0.05 was considered significant for the presence of horizontal pleiotropy (29). The Cochran’s Q test was used to assess heterogeneity, and P < 0.05 was considered significant for the presence of heterogeneity (33). The leave-one-out plots were used to assess the influence of individual variants on observed associations (34). The MR Steiger directionality tests were used to assess whether there was reverse causality in our *cis*-MR study (35). Reverse MR aimed to evaluate whether MP was causally associated with sVEGFR2 or sVEGFR3 levels and was employed using all the aforementioned methods. Unavailable variants in MP were omitted and no proxy variants were used to substitute them.

### 2.4 Bayesian colocalization analyses

Bayesian colocalization was performed to investigate whether sVEGFR2 or sVEGFR3 and MP shared genetic variants within ±100 kb around the *VEGFR2* or *VEGFR3* gene, respectively. The “coloc” package (version 5.2.3) in R (version 4.4.1) was utilized for these analyses (36), with visualization facilitated by the “locuscomparer” package (version 1.0.0) (37). The posterior probability of hypothesis 4 (PPH_4_) > 0.8 indicated strong evidence of colocalization.

### 2.5 Ethical statement

In this study, we utilized publicly available GWAS data and ethical approvals were obtained in all original studies.

### 2.6 Statistical analysis

The IVW method was primarily used to assess the causal effect. MR results were reported as odds ratios (OR) with 95% confidence interval (CI) per one-SD increase in log10 ng/ml of sVEGFR2 (or others) levels. The statistical power was calculated in “mRnd” (https://shiny.cnsgenomics.com/mRnd/) (38). All MR analyses were two-sided and performed using package ‘TwoSampleMR’ (version 0.6.4) in R (version 4.4.1). P < 0.05 was considered statistically significant.

## 3 Results

We selected genome-wide genetic variants associated with levels of sVEGFR2, sVEGFR3, VEGF-A, or VEGF-D, as well as gene-restricted genetic variants associated with levels of sVEGFR2, sVEGFR3, VEGF-A, or VEGF-C. These variants were utilized for univariable MR and *cis*-MR analyses, respectively, to evaluate the effects of genetically predicted levels of each molecule on MP using data from the FinnGen study. All genetic variants demonstrated strong validity (F-statistic > 10) (Supplementary table 1-10). None of these genetic variants showed significant associations with MP in our study.

Genetically predicted higher levels of sVEGFR2 demonstrated lower odds of MP in both univariable MR (OR 0.89, 95% CI 0.81-0.97, P=8.93 × 10^-3^, statistical power 0.73) and *cis*-MR (OR 0.82, 95% CI 0.75-0.89, P = 9.20 × 10^-6^, statistical power 0.87) (Table 1 and 2). However, while *cis*-MR suggested that higher sVEGFR3 levels were associated with lower odds of MP (OR 0.71, 95% CI 0.57-0.89, P=2.64 × 10^-3^, statistical power 0.79) (Table 2), this finding was not consistent with the results from unviable MR (OR 0.95, 95% CI 0.88-1.02, P=0.154) (Table 1). Univariable MR and/or *cis*-MR suggested no associations between VEGF-A, VEGF-C, or VEGF-D levels and MP (Table 1-2 and Supplementary table 11-12). Statistical power calculations for the significant MR results were presented in Supplementary table 13.

**Table 1.** Univariable MR results for the relationship between VEGF family and MP.

| Method | # of SNPs | OR (95% CI) | P |
| --- | --- | --- | --- |
| Relative sVEGFR2 levels on MP |  |  |  |
| IVW | 19 | 0.89 (0.81-0.97) | $8.93 \times 10^{-3}$ |
| Weighted median | | 0.85 (0.77-0.95) | $2.45 \times 10^{-3}$ |
| MR-Egger | | 0.81 (0.71-0.93) | $6.28 \times 10^{-3}$ |
| Relative sVEGFR3 levels on MP |  |  |  |
| IVW | 26 | 0.95 (0.88-1.02) | 0.154 |
| Weighted median |  | 0.93 (0.84-1.02) | 0.119 |
| MR-Egger |  | 0.95 (0.85-1.06) | 0.395 |
| Relative VEGF-A levels on MP |  |  |  |
| IVW | 3 | 0.94 (0.82-1.09) | 0.434 |
| Weighted median |  | 0.94 (0.81-1.09) | 0.403 |
| MR-Egger |  | 0.90 (0.70-1.16) | 0.569 |
| Relative VEGF-D levels on MP |  |  |  |
| IVW | 5 | 1.13 (0.84-1.54) | 0.417 |
| Weighted median |  | 1.16 (0.82-1.66) | 0.402 |
| MR-Egger |  | 1.39 (0.60-3.26) | 0.499 |

MP, macular pucker; sVEGFR2/3, soluble vascular endothelial growth factor receptor 2/3; #, number; SNP, single nucleotide polymorphism; IVW, inverse-variance weighted; all statistical tests were two-sided; results were reported as odds ratio (OR) with 95% confidence interval (CI) per one-standard deviation increase in log<sub>10</sub> ng/ml of indicated protein levels; P value < 0.05 was considered significant.

**Table 2.** *Cis*-MR results for the relationship between VEGF family and MP.

| Method | # of SNPs | OR (95% CI) | P | Cochran's Q-P | MR-Egger intercept-P | MR-PRESSO global test-P |
| --- | --- | --- | --- | --- | --- | --- |
| **Relative sVEGFR2 levels on MP** |  |  |  |  |  |  |
| IVW | | 0.82 (0.75-0.89) | $9.20 \times 10^{-6}$ | | 0.563 | |
| Weighted median | 17 | 0.78 (0.69-0.88) | $8.79 \times 10^{-5}$ | 0.586 | - | 0.547 |
| MR-Egger | | 0.80 (0.71-0.90) | $2.05 \times 10^{-3}$ | | - | |
| **Relative sVEGFR3 levels on MP** |  |  |  |  |  |  |
| IVW | | 0.71 (0.57-0.89) | $2.64 \times 10^{-3}$ | | 0.47 | |
| Weighted median | 8 | 0.75 (0.57-0.98) | $3.64 \times 10^{-2}$ | 0.881 | - | 0.844 |
| MR-Egger |  | 0.86 (0.51-1.45) | 0.592 |  | - |  |
| **Relative VEGF-A levels on MP** |  |  |  |  |  |  |
| IVW |  | 1.01 (0.89-1.14) | 0.936 |  | 0.891 |  |
| Weighted median | 13 | 0.94 (0.82-1.08) | 0.373 | 0.244 | - | 0.452 |
| MR-Egger |  | 0.99 (0.73-1.32) | 0.929 |  | - |  |
| **Relative VEGF-C levels on MP** |  |  |  |  |  |  |
| IVW |  | 0.99 (0.86-1.15) | 0.941 |  | 0.899 |  |
| Weighted median | 3 | 0.97 (0.82-1.16) | 0.777 | 0.637 | - | - |
| MR-Egger |  | 1.13 (0.24-5.25) | 0.904 |  | - |  |

MP, macular pucker; sVEGFR2/3, soluble vascular endothelial growth factor receptor 2/3; #, number; SNP, single nucleotide polymorphism; IVW, inverse-variance weighted; MR-PRESSO, Mendelian Randomization Pleiotropy RESidual Sum and Outlier; -, not applicable; all statistical tests were two-sided; results were reported as odds ratio (OR) with 95% confidence interval (CI) per one-standard deviation increase in log10 ng/ml of indicated protein levels; P value < 0.05 was considered significant.

Scatter plots showing the relationship between levels of sVEGFR2 or sVEGFR3 and MP for the genetic variants were presented in Figure 1, with colored lines representing the slopes from different regression analyses of *cis*-MR (Figure 1 and Supplementary table 12). *Cis*-MR forest plots showing the effects of genetic variants that were associated with sVEGFR2 or sVEGFR3 levels on MP were presented in Supplementary figure 1a and 1b. The leave-one-out plots in *cis*-MR revealed that no individual genetic variant significantly influenced the overall outcomes, suggesting a consistent pattern of inverse associations between levels of sVEGFR2 or sVEGFR3 (potentially) and MP (Supplementary figure 2a and 2b). Across univariable MR and *cis*-MR analyses, the MR-Egger intercept did not indicate evidence of horizontal pleiotropy, and MR-PRESSO did not identify any potential outliers (Table 2 and Supplementary table 11). Cochran’s Q statistic also indicated no evidence of heterogeneity (Table 2 and Supplementary table 11). Moreover, MR Steiger directionality tests and reverse MR analyses suggested no evidence of reverse causality in the *cis*-MR results (Table 3).

**Figure 1.**
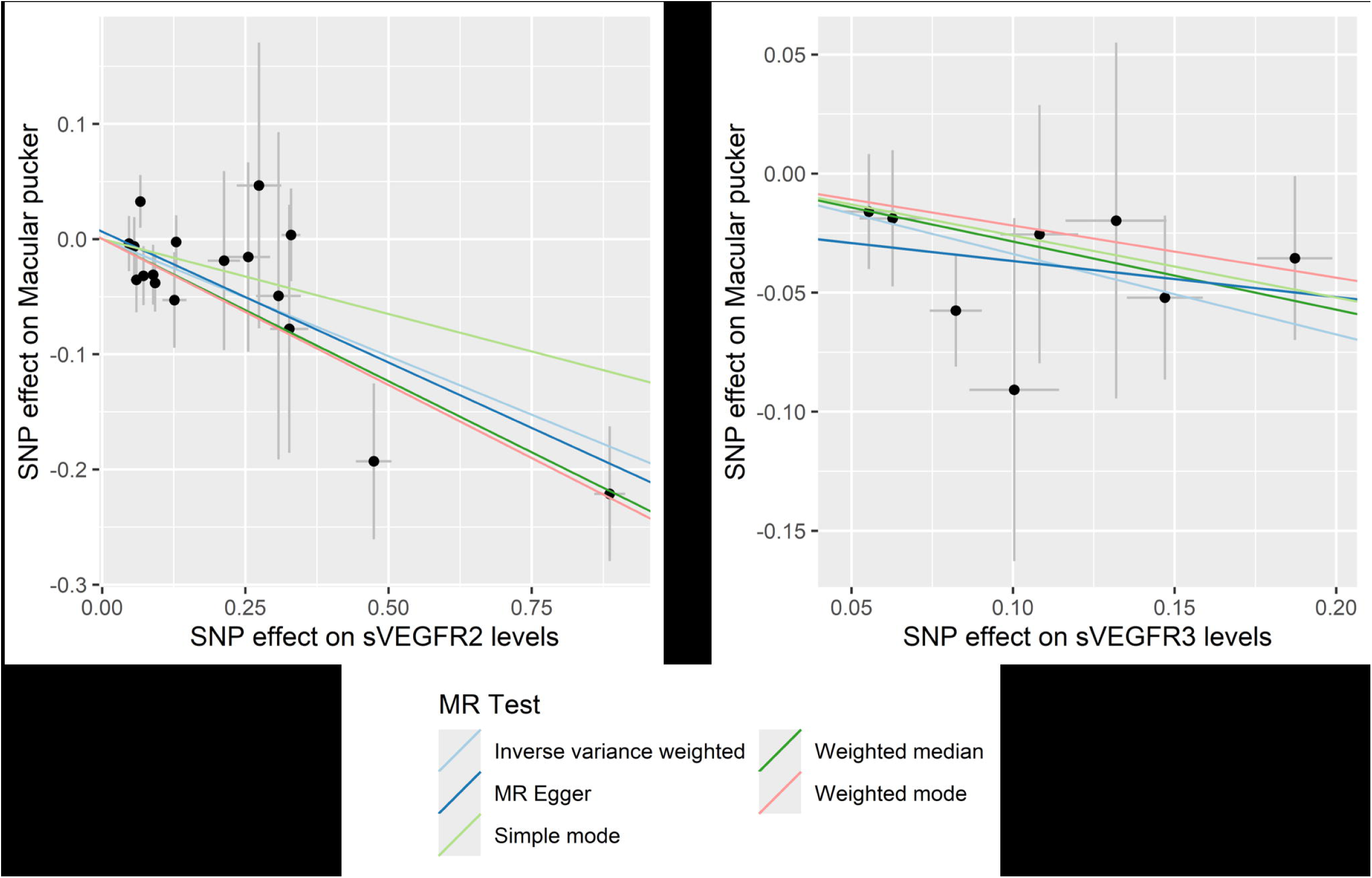
*Cis*-MR scatter plots for the relationship of sVEGFR2 or sVEGFR3 levels with Macular pucker (MP). The x-axis shows the effect of each SNP on sVEGFR2 levels (a) or sVEGFR3 levels (b). The y-axis shows the effect of each SNP on MP. The regression lines for inverse variance weighted (multiplicative random effects), weighted median, MR-egger, simple mode, weighted mode method are shown. The data are presented as raw β values with corresponding 95% confidence intervals. sVEGFR2 and sVEGFR3 protein quantitative trait loci of genome-wide association studies (GWAS) were from deCODE genetics, N=35,559 samples; MP GWAS was from the FinnGen study, N=3,974 cases and N=376,650 controls; SNP, Single nucleotide polymorphisms; sVEGFR2/3, soluble vascular endothelial growth factor receptor 2/3.

**Table 3.** Reverse causality tests.

| **MR Steiger directionality tests for the causality in *cis*-MR results in table 2** |  |  |  |  |  |
| --- | --- | --- | --- | --- | --- |
| exposure | outcome | snp_r2.exposure | snp_r2.outcome | correct_causal_direction | steiger_Pval |
| sVEGFR2 levels | MP | 9.93E-02 | 6.21E-05 | TRUE | 1.80E-118 |
| sVEGFR3 levels | MP | 2.28E-02 | 3.17E-05 | TRUE | 4.44E-153 |
| **Reverse MR results of the IVW regression** |  |  |  |  |  |
| exposure | outcome | # of SNPs | OR (95% CI) | P |  |
| MP | sVEGFR2 levels | 3 | 1.04 (0.98-1.10) | 0.209 |  |
| MP | sVEGFR3 levels | 3 | 1.00 (0.90-1.12) | 0.939 |  |

sVEGFR2/3, soluble vascular endothelial growth factor receptor 2/3; MP, macular pucker; #, number; SNP, single nucleotide polymorphism; MR, mendelian randomization; IVW, inverse-variance weighted; OR, odds ratio; CI, confidence interval; all statistics were two-sided and P < 0.05 was considered significant.

Bayesian colocalization analyses were performed to further validate the significant associations found in the *cis*-MR. These analyses indicated that the *VEGFR2* gene region was shared between sVEGFR2 and MP (PPH_4_ = 0.94) (Figure 2a). Conversely, no shared causal variants were evidenced in the *VEGFR3* gene region between sVEGFR3 and MP (PPH_4_= 0.03) (Figure 2b).

**Figure 2.**
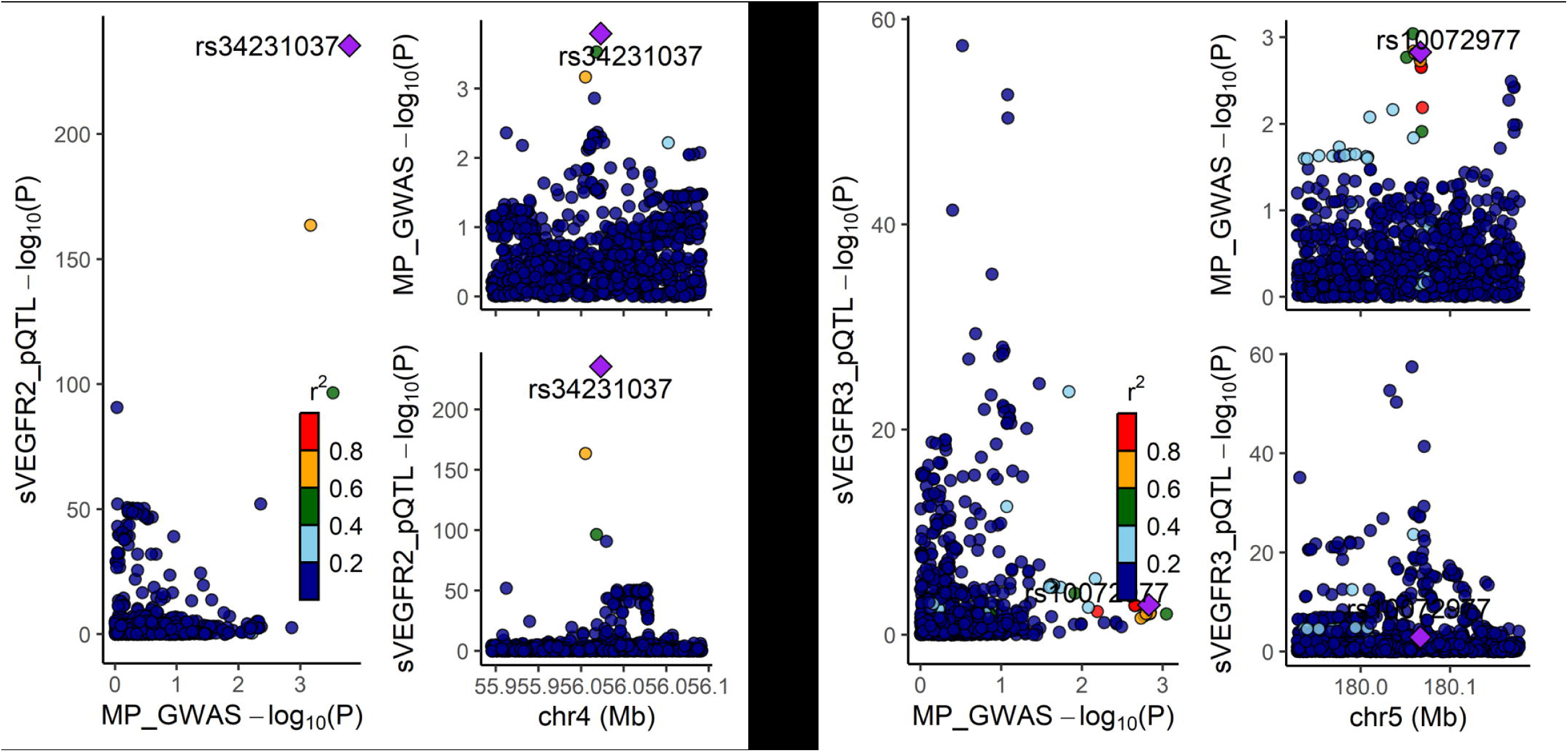
Colocalization between sVEGFR2 or sVEGFR3 and Macular pucker (MP). Bayesian colocalization analysis was conducted to investigate whether (a) sVEGFR2 or (b) sVEGFR3 and MP share genetic variants within ±100 kb around the *VEGFR2* gene (chromosome 4, position 55,078,481-55,125,595) or *VEGFR3* gene (chromosome 5, position 180,601,506-180,650,298), respectively. A posterior probability of hypothesis 4 (PPH_4_) > 0.8 indicated strong evidence of colocalization. In the regional plots, each dot represented a genetic variant, and the candidate causal variant was depicted as a purple diamond. The color of other variants indicated their linkage disequilibrium (r²). sVEGFR2 and sVEGFR3 protein quantitative trait loci of genome-wide association studies (GWAS) were from deCODE genetics, N=35,559 samples; MP GWAS was from the FinnGen study, N=3,974 cases and N=376,650 controls; sVEGFR2/3, soluble vascular endothelial growth factor receptor 2/3.

In summary, integrating insights from univariable MR, *cis*-MR, and colocalization analyses suggested that the protective causal effects of sVEGFR2 on MP should warrant stronger consideration compared to those of sVEGFR3.

## 4 Discussion

To the best of our knowledge, our *cis*-MR and colocalization study provided the first human causal evidence that higher sVEGFR2 levels were associated with a reduced risk of MP. Therefore, these analyses proposed sVEGFR2 as a promising target for preventing or treating MP.

Initially, we conducted univariable MR to explore potential causal effects of several members within the VEGF/VEGFR family on MP (Figure 1 and Supplementary table 11). We utilized genome-wide genetic variants selected under stringent LD-clumping criteria, with parallel gene-restricted variants used for *cis*-MR as discussed below (Methods and Supplementary table 1). VEGF-C was excluded from univariable MR due to the unavailability of associated variants meeting these criteria. Additionally, sVEGFR1 and VEGF-B were omitted from all analyses due to the unavailability of high-quality GWAS data (24). Univariable MR analyses indicated that elevated sVEGFR2 levels were associated with a decreased risk of MP. In contrast, no significant associations were observed for other examined members of the VEGF/VEGFR family by univariable MR, including sVEGFR3, VEGF-A, and VEGF-D.

Subsequently, we utilized *cis*-MR, a method leveraging gene-restricted variants to minimize potential pleiotropy and reverse causality, to evaluate the causal relationship between the VEGF/VEGFR family and MP. VEGF-D was excluded from analysis due to its gene’s location on the X chromosome. Again, elevated levels of sVEGFR2 were associated with decreased MP risk in *cis*-MR analyses, with a statistical power of 0.87. In contrast to findings from univariable MR, *cis*-MR indicated an inverse association between sVEGFR3 levels and MP risk (OR 0.71, 95% CI 0.57-0.89, P=2.64 × 10^-3^, statistical power 0.79). Rigorous sensitivity tests indicated no evidence of pleiotropy (MR-Egger intercept and MR-PRESSO global test), reverse causality (MR-Steiger and reverse MR), or heterogeneity (Cochran’s Q), supporting the robustness of our *cis*-MR results (34). The discrepant findings regarding sVEGFR3’s impact on MP in univariable MR versus *cis*-MR analyses likely reflected its role as a mediator of MP. In addition, colocalization analysis suggested no shared genetic variants in the *VEGFR3* gene region between sVEGFR3 and MP (PPH_4_=0.03), supporting the hypothesis that sVEGFR3 may mediate rather than directly affect MP. Conversely, colocalization analyses revealed shared genetic variants in the *VEGFR2* gene region between sVEGFR2 and MP (PPH_4_=0.94), thereby strengthening the evidence for causally protective role of sVEGFR2 in MP as identified both in univariable MR and in *cis*-MR.

VEGFR2 is expressed in both retinal pigment epithelium (RPE) and endothelial cells, with endothelial cells being more prevalent in vascular MP (8; 10). VEGF-A is primarily expressed by RPEs and is prominent in both vascular and avascular MP (8; 10). These characteristics suggest that the pathogenesis of MP involves complex interactions among diverse cellular responses and the VEGF-A/VEGFR2 signaling at the vitreoretinal interface (3; 5; 6; 7). Therefore, the protective role of sVEGFR2 in MP likely encompasses multiple biological mechanisms. As a decoy receptor, sVEGFR2 sequesters VEGF-A and/or VEGF-C, competing for binding with mVEGFR2 and/or mVEGFR3, respectively (13; 14; 15; 16; 17; 18). It could thereby regulate downstream angiogenesis, lymphangiogenesis, inflammation, and fibrotic responses in the macula (6; 10; 14), potentially preventing or alleviating the development of MP. Sequestering VEGF-A by sVEGFR2 could also prevent VEGF-A/mVEGFR2-mediated vascular permeability (11; 12), thereby stabilizing retinal vasculature and potentially mitigating MP pathogenesis. Continued research is needed to fully understand sVEGFR2’s protective role in MP and its therapeutic potential.

In addition, our univariable MR and *cis*-MR analyses indicated no significant associations between VEGF-A, VEGF-C, or VEGF-D and MP. This lack of association may be explained by their tightly regulated functions through biological mechanisms such as sVEGFRs acting as competitive decoys against mVEGFRs (14; 15; 39), potentially diminishing the effects of VEGF/mVEGFR signaling on MP, as observed in our analyses.

Our study showing causally protective effects of sVEGFR2 on MP through *cis*-MR and colocalization had several major strengths. We leveraged genetic associations from a large GWAS of pQTL, which could maximize the inclusion of genetic variants associated with levels of interest members in the VEGF/VEGFR family and enhance statistical power (21). Additionally, genetic variants were selected from regions proximal to the regions of the respective gene. These variants were likely to have a significant impact on protein expression compared to their impact on other traits. Therefore, *cis*-MR was less likely to violate the assumption of ‘no horizontal pleiotropy’ (22). Besides, utilizing *cis*-MR for protein risk factors could enhance the reliability of causal inferences by aligning with Crick’s central dogma. This principle could support a sequential flow from gene to protein to disease, which could be more plausible than a gene to disease to protein pathway, particularly in studies using population-based samples. Therefore, *cis*-MR could effectively minimize the likelihood of reverse causality (22). Furthermore, Bayesian colocalization analysis is a powerful method used to determine whether two different traits or genomic signals are influenced by the same causal genetic variants. Integrating *cis*-MR with colocalization could synergistically harness their respective strengths (21; 22; 23), reinforcing the causal relationship between sVEGFR2 and MP.

However, our study had limitations. While we identified genetic variants significantly associated with sVEGFR2 levels, they explained only a small proportion of the total variance and should not be considered as exact proxies of the exposure (21). Moreover, MR effect estimates assume lifelong exposure to altered protein levels (e.g., sVEGFR2), which could potentially be diverging from observational associations and therapeutic interventions (21). Besides, sVEGFR2 levels measured in plasma from the published GWAS pQTL analysis may not accurately represent their concentrations or biological functions in retinal tissues (24), where they could potentially play a pivotal role in MP pathogenesis. This indicated the need for cautious interpretation of our findings. Our study primarily highlighted the causally protective effects of sVEGFR2 on MP, yet the underlying biological mechanisms require further investigation. Lastly, our study included only individuals of European descent (24; 25), which could limit the generalizability to other populations.

## 5 Conclusion

In conclusion, our study employed large GWAS data and utilized *cis*-MR and colocalization analyses to provide robust genetic evidence supporting a protective effect of sVEGFR2 against MP. These findings suggested sVEGFR2 as a promising drug target for MP and warrant future basic and translational research in this area.

## Data Availability

All GWAS data are publicly available.

https://www.decode.com/summarydata/

https://www.finngen.fi/en/access_results

## 6 Author contributions

Study design: J.G., Q.G.; data acquisition and analysis: J.G., Q.G.; figures, tables and original draft: J.G.; writing, reviewing and editing: J.G., Q.G.

## 8 Data Sharing Statement

All GWAS data are publicly available. The pQTL GWAS was obtained from deCODE genetics (https://www.decode.com/summarydata/). The MP GWAS was obtained from the FinnGen study (https://www.finngen.fi/en/access_results).

## 9 Acknowledgements

We gratefully acknowledge the participants and investigators of the FinnGen study and deCODE genetics for generously sharing the GWAS data, which has been instrumental in our research. We sincerely thank our colleagues at Department of Cardiovascular Diseases, the First Hospital of Jilin University, for their insightful discussions.

## 10 Supplementary materials

Supplementary materials associated with this article can be found in the online version, at xxx.

---

**Supplementary figure 1.**
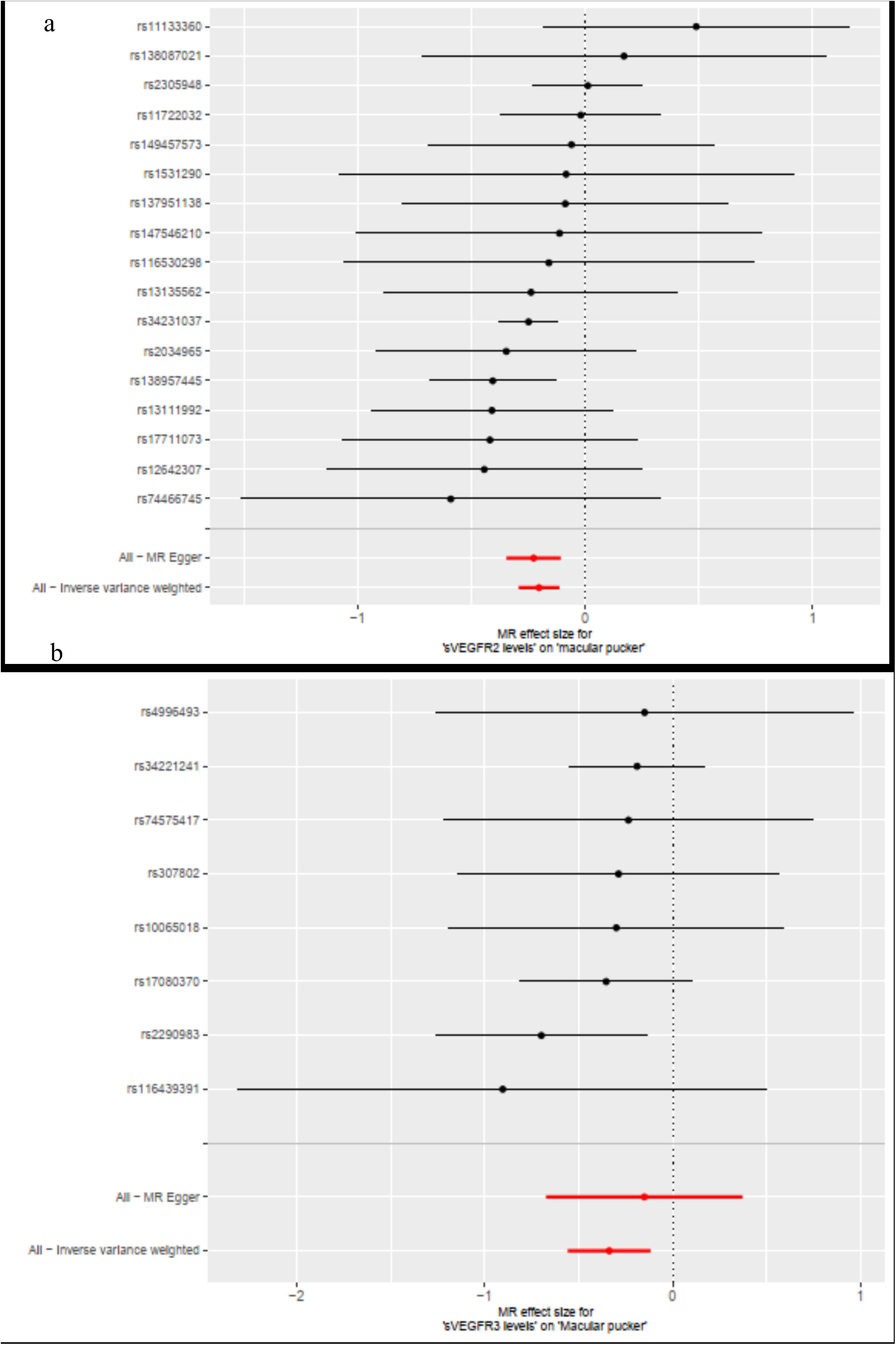
*Cis*-MR forest plots for the relationship of sVEGFR2 or sVEGFR3 levels with Macular pucker (MP). The x-axis shows the *cis*-Mendelian Randomization (MR) effect size for (a) sVEGFR2 levels or (b) sVEGFR3 levels on MP. The y-axis shows the analysis for each of the SNPs and for the SNPs in total using the MR-egger and inverse variance weighted (multiplicative random effects) methods. The data are presented as raw β values with corresponding 95% confidence intervals. sVEGFR2 and sVEGFR3 protein quantitative trait loci of genome-wide association studies (GWAS) were from deCODE genetics, N=35,559 samples; MP GWAS was from the FinnGen study, N=3,974 cases and N=376,650 controls; SNP, Single nucleotide polymorphisms; sVEGFR2/3, soluble vascular endothelial growth factor receptor 2/3.

**Supplementary figure 2.**
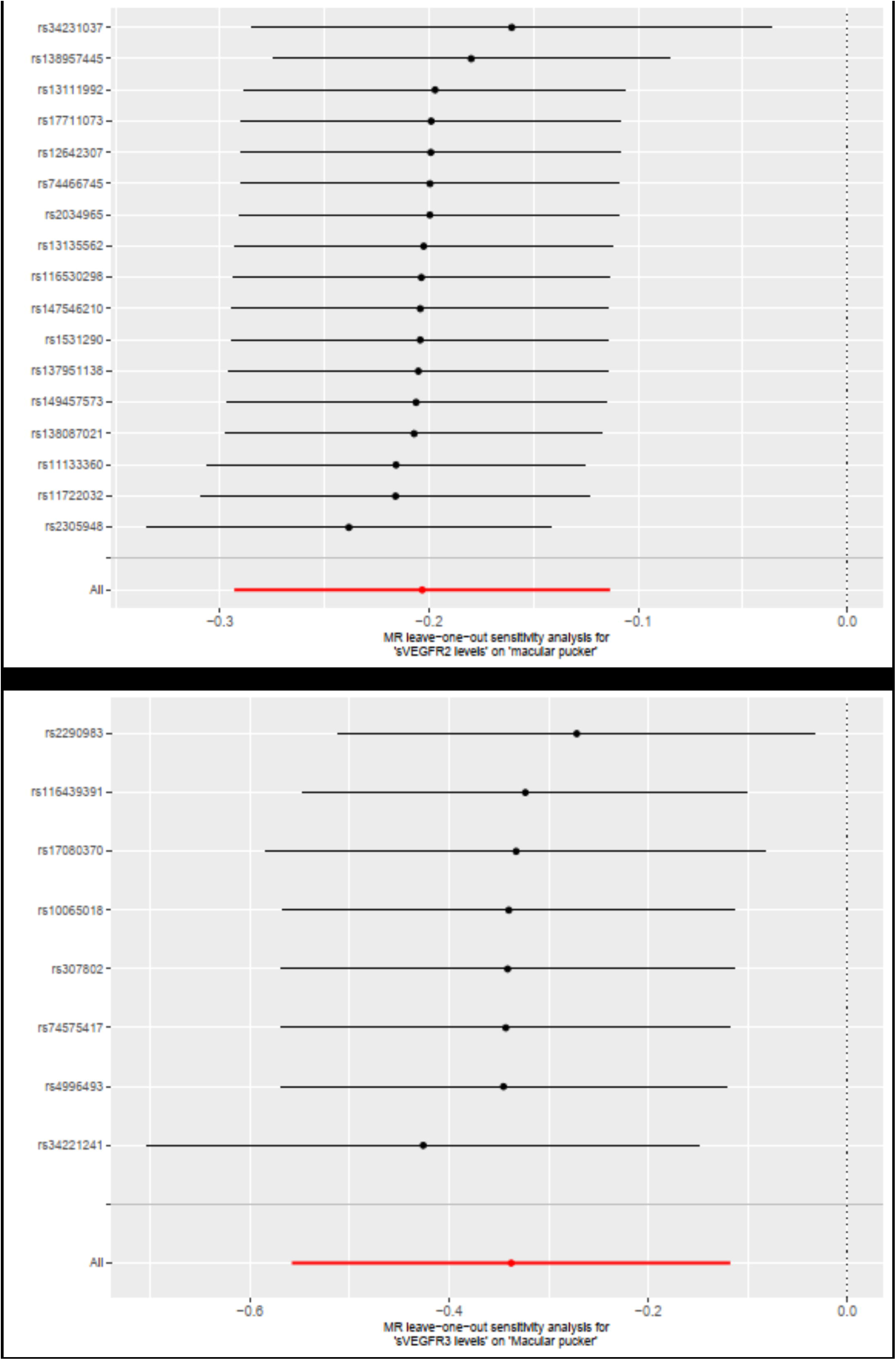
*Cis*-MR leave-one-out plots for the effect of sVEGFR2 or sVEGFR3 levels on Macular pucker (MP). The x-axis shows the *cis*-Mendelian Randomization (MR) leave-one-out sensitivity analysis for the effect of (a) sVEGFR2 levels or (b) sVEGFR3 levels on MP. The y-axis shows the analysis for leave-one-out of SNPs and the effect of the total SNPs on MP. The data are presented as raw β values with corresponding 95% confidence intervals. sVEGFR2 and sVEGFR3 protein quantitative trait loci of genome-wide association studies (GWAS) were from deCODE genetics, N=35,559 samples; MP GWAS was from the FinnGen study, N=3,974 cases and N=376,650 controls; SNP, Single nucleotide polymorphisms; sVEGFR2/3, soluble vascular endothelial growth factor receptor 2/3.

